# Remote Ischemic Conditioning for Neonatal Hypoxic-Ischemic Encephalopathy: A Safety and Feasibility Trial

**DOI:** 10.1101/2024.05.17.24307297

**Authors:** Emily Lo, Mehmet N. Cizmeci, Diane Wilson, Linh G. Ly, Amr El-Shahed, Martin Offringa, Agostino Pierro, Brian T. Kalish

## Abstract

**Objective:** To test the hypothesis that remote ischemic conditioning (RIC) is safe and feasible as an adjunctive neuroprotective treatment in neonates with hypoxic-ischemic encephalopathy (HIE) during therapeutic hypothermia (TH).

**Study design:** In this prospective, randomized, safety and dose escalation study in 32 neonates with HIE undergoing TH at a single quaternary referral NICU, four cohorts of consecutive patients received escalating therapy as follows: the first cohort of four patients received 3 minutes of limb ischemia by inflating a blood pressure cuff followed by 3 minutes of reperfusion; the second cohort of four patients received 5 minutes of limb ischemia followed by 5 minutes of reperfusion; the third cohort of four patients received 5 minutes of limb ischemia followed by 5 minutes of reperfusion on days 1 and 2 of TH; and the last cohort of four patients received 5 minutes of limb ischemia followed by 5 minutes of reperfusion on days 1, 2, and 3 of TH. For patients randomized to the control arm (n=16), a blood pressure cuff was applied without inflation. Each patient received 4 cycles of RIC or sham. Clinical, biochemical, and safety outcomes were monitored in both groups.

**Results:** All patients received the designated RIC therapy without interruption or delay on days 1-3 of TH. RIC was not associated with increased pain, vascular, cutaneous, muscular, or neural safety events. There was no difference in the incidence of seizures, brain injury, or mortality between the two groups with the escalation of RIC dose and frequency.

**Conclusion:** RIC is a safe and feasible adjunctive therapy for neonates with HIE undergoing TH. Future studies to investigate the potential efficacy of RIC for HIE are warranted.

## INTRODUCTION

Hypoxic-ischemic encephalopathy (HIE) is a potentially devastating neurological condition in neonates that results from a peripartum disruption in blood flow and oxygen delivery to the fetus. HIE is estimated to affect 1.5 per 1000 live births worldwide, making it a leading cause of neonatal morbidity and mortality (1). Despite the implementation of therapeutic hypothermia (TH) as the standard of care for infants with moderate-severe HIE, approximately 50% of infants with HIE die or have neurodevelopmental impairment (2). Survivors have increased risk of seizure disorders, hearing loss, and vision loss (3). Thus, novel adjunctive strategies are urgently needed to provide neuroprotection after HIE.

Here, we propose remote ischemic conditioning (RIC) as a potential neuroprotective adjunct in infants with HIE. RIC is a maneuver that creates transient cycles of ischemia and reperfusion to a remote site (e.g. limb), typically through extended and repeated inflation of a standard blood pressure cuff. The goal is to protect the organ exposed to ischemia, such as the brain, from further injury by inducing non-lethal ischemia in the remote site. Clinical trials in adults with acute ischemic stroke have found that RIC was associated a reduction in the area of infarct and the incidence of recurrent strokes, as well as with improved neurological outcomes, albeit with heterogeneous effects, likely secondary to patient selection and RIC implementation (4–6). This is consistent with preclinical rodent studies of stroke where RIC significantly reduced the extent of ischemia and neurological deficits (7). Furthermore, RIC was found to be safe and feasible in neonates of younger gestational age and lower birth weight (8). However, the effects of RIC on neonates with HIE undergoing TH has not been explored. We propose that RIC may be a promising adjunctive therapeutic option to improve neurodevelopmental outcomes. Therefore, the objective of this study was to assess the safety and feasibility of RIC in neonates with HIE.

## METHODS

This was a single-site, single-blinded, prospective, and randomized controlled trial of RIC in neonates with HIE undergoing TH. Patients were enrolled between January 2022 to January 2024. The trial was approved by the Research Ethics Board (REB) at The Hospital of Sick Children (REB #1000077295) and registered with ClinicalTrials.gov (NCT05379218). Informed consent was obtained from parent or legal guardian for all participants.

### Participants

Neonates who were admitted to the NICU with a diagnosis of HIE and who qualified for TH based on the attending neonatologist’s assessment were eligible for enrollment. The flow diagram of patients in the study is shown in Figure 1. In the current TH protocol for HIE at the Hospital for Sick Children, infants who qualify for TH should fulfill the following criteria: gestational age greater than or equal to 35 weeks, less than 6 hours of life, evidence of intrapartum hypoxia defined as either cord or postnatal gas within one hour of birth with pH ≤ 7 or base deficits ≥ -16 or if pH 7.01-7.15 or base deficit -10 to -15 with acute perinatal event, and signs of moderate to severe encephalopathy or seizures; he ultimate decision to proceed with TH is based on the neonatologist’s discretion. Exclusion criteria were as follows: infants with gestational age <35 weeks, known central nervous system malformations, known chromosomal or genetic anomalies, confirmed or suspected inborn errors of metabolism, parental decision for withdrawal, patients requiring significant hemodynamic support (two or more blood pressure support agents, epinephrine infusion >0.05 mcg/kg/min, or vasopressin >0.1 mU/kg/min) for the four hour period prior to RIC, and infants requiring inhaled nitric oxide or fraction of inspired oxygen (FiO2) >0.50 for the four hour period prior to RIC.

### Randomization and dose escalation

Patients were randomized and allocated to the RIC intervention or control arm by sequentially numbered, opaque, and sealed envelopes. For patients randomized to the RIC arm, cohorts of four consecutive patients received escalating therapy:

A. Four consecutive patients had four cycles of three minutes of ischemia, followed by five minutes of reperfusion on day 1 of TH.
B. Observing no safety events from the patients in group (A), four consecutive patients had four cycles of five minutes of ischemia, followed by five minutes of reperfusion on day 1 of TH.
C. Observing no safety events from patients in group (B), four consecutive patients had four cycles of five minutes of ischemia, followed by five minutes of reperfusion on day 1 and 2 of TH.
D. Observing no safety events from patients in group (C), four consecutive patients had four cycles of five minutes of ischemia, followed by five minutes of reperfusion on day 1, 2, and 3 of TH.

Enrolled patients had the first RIC or sham procedure performed within 24 hours of TH initiation.

### Procedure

For patients randomized to the RIC intervention arm, an appropriately sized manual blood pressure cuff and a pulse oximeter probe were applied on an arm or leg and inflated and deflated to administer ischemia-reperfusion cycles. The limb selected for RIC was free from intravenous or intra-arterial catheters. Cycles of ischemia and reperfusion were achieved with the inflation and deflation of a manual blood pressure cuff. This cuff was inflated 20 mm Hg above the participants’ systolic blood pressure as measured by an arterial line and/or manual blood pressure cuff. To ensure that adequate ischemia was achieved, loss of the pulse oximeter probe signal was required. If loss of pulse oximeter probe signal was not achieved, the inflation pressure was increased by 5 mm Hg every 10 seconds until loss of pulse oximeter probe signal. The timing of inflation and deflation was dependent on the cohort group (A-D), as described above. For patients randomized to the control arm, a blood pressure cuff and pulse oximeter probe were applied to a limb, but the cuff was not inflated.

### Primary Outcome: Feasibility and Safety Assessments

Intervention feasibility was defined as the ability to complete RIC in a dose escalation therapy in neonates undergoing TH. Several safety parameters, including pain, vascular, cutaneous, muscular, and neural components, were assessed prior, during, and after the ischemia-reperfusion cycles. Pain was measured by the Premature Infant Pain Profile (PIPP) score 15 minutes prior to the first cycle, 30-60 seconds after each cycle, and 15 minutes and 6 hours after the last cycle. Vascular limb components were assessed by perfusion (capillary refill time) and return of pulse oximetry wave 60-90 seconds after every cuff deflation. Cutaneous limb components were assessed by visual inspection of the skin for wounds, skin excoriations, petechiae, and/or hematomas after every cycle and 24 hours after the last cycle of ischemia-reperfusion. Muscular limb components were assessed through laboratory markers of potassium, phosphate, and creatinine as muscle ischemia can result in metabolic (hyperkalemia, hyperphosphatemia) and renal disturbances. Laboratory measurements were done at 24, 48, and 72 hours of TH as per the TH protocol. Laboratory measurements were collected +/- 8 hours from the 24, 48, and 72 hours time points. If there was missing data at the 72-hour time point, the 48-hour laboratory values were collected and used for statistical analysis. Neural limb components were assessed by eliciting grasp reflexes before the first cycle, immediately after RIC, and 24 hours after the last RIC cycle.

### Criteria for acute RIC Interruption and Rescue Interventions

The maneuver was stopped if the arm/leg undergoing RIC showed absence of return of perfusion by clinical inspection or absence pulse return by pulse oximetry reading within the reperfusion time of 5 minutes. If either of these events occurred, no additional RIC cycles were applied to the patient. If the perfusion does not return after the expected reperfusion time, escalation of rescue interventions include: rewarming the limb for 5 minutes, elevating the limb for 5 minutes, applying a nitroglycerine patch over the skin area covered by the cuff, requesting for a doppler ultrasound, and requesting for a thrombosis consult.

### Brain MRI Assessments

All newborns underwent brain MRI as part of the routine clinical care that followed the institutional guidelines. The MRI scans were acquired on 3 Tesla (Magnetom Skyra, Siemens Healthcare Limited, Germany) scanners with an age-appropriate head coil. The 3D sagittal T1-weighted images were acquired with a slice-thickness of 0.5 mm, resolution 0.4018 mm x 0.4018 mm, echo time 3.52 ms, repetition time 2200 ms, while the 2D T2-weighted volumes were acquired with a slice thickness of 3 mm, in-plane resolution 0.5 mm x 0.5 mm, echo time 186 ms and the repetition time 5330 ms. The aimed timing of the MRI scans was on days 4 to 5 after birth, after the completion of the rewarming phase of TH. An experienced reviewer officially trained in neonatal neurology and neuroimaging (MNC) who was blinded to the clinical data systematically assessed and scored the MRI scans for the pattern of WM and DGM abnormalities by using a validated brain injury scoring system developed by Weeke et al (9). The cumulative score for DGM, MR-spectroscopy metrics, WM and cortical regions, cerebellum, and additional findings on brain MRI was calculated to obtain a total brain injury score ranging from 0 to 55 points, with higher scores reflecting greater severity of brain abnormalities.

### Statistical Methods

Statistical analyses were performed on R (version 4.3.2). An unpaired two-tailed Student’s t-test or Wilcoxon rank-sum test was used to compare continuous variables and a chi-square test or Fisher’s exact test was used to compare categorical variables between the control and RIC groups. Continuous variables were plotted and tested for normality. Continuous variables were presented as mean (± standard deviation [SD]). Categorical variables were presented as numbers and percentages. P value <0.05 was considered statistically significant. P value <0.05 was considered statistically significant.

### Data Safety and Monitoring

A Data Safety and Monitoring Board (DSMB) consisting of pediatric cardiologists and critical care intensivists convened 4 times during the clinical trial to review the interim safety data on all patients after completion of each block of 8 patients. Enrollment was paused until the DSMB determined it was safe and feasible to proceed to the next dose cohort.

## RESULTS

Out of 114 eligible patients, thirty-two neonates were recruited in this study in a blocked treatment assignment of 8 patients (4 in RIC and 4 in control) in each cohort (Figure 1). The baseline and clinical characteristics of these patients are shown in Table 1. There was no significant difference in gestational age (39 ± 1.2 versus 39.5 ± 1.4 weeks), birth weight (3416 ± 701 versus 3617 ± 525 grams), and perinatal variables, including Apgar scores and cord gases, between the control and RIC groups. The Modified Sarnat scores on admission were not different between the two groups.

### Primary Outcomes: RIC Feasibility and Safety

RIC cycles were administered as planned, including within 24 hours of TH initiation, in all neonates. The clinical outcomes between the control and RIC groups are shown in Table 2. There were no incidences of absence of return of perfusion, RIC interruption and need for rescue intervention in either the control or RIC groups. While there were two incidences of subcutaneous fat necrosis and two incidences of AKI (stage 2 and stage 3) in the control group, there were no incidences in the RIC group. The incidence of seizures (clinical or subclinical) was comparable between the control and RIC groups (10 versus 9 infants, p=0.296). Similarly, mortality was comparable between the control and RIC groups (4 versus 5 infants, p=1.00).

### Secondary Outcomes

There were no incidences of cutaneous injury, including skin breakdown, bruising, ecchymosis, or petechiae, or neural injury, including loss of grasp reflexes, in either group. There was also no difference in need for analgesia after RIC or sham (1 in each arm).

The physiological changes during RIC or sham are shown in Table 4; these clinical parameters are displayed in Figures 2-3. There was no statistical difference in oxygen saturation, capillary refill time, and PIPP scores pre- and post-maneuver between the control and RIC groups across all cohorts. Similarly, there was no statistical significance in the biochemical markers between the control and RIC group at 24 hours and at 48 to 72 hours, with the exception of creatinine which was higher in the control group compared to the RIC group at 24 hours (83 ± 31 versus 61 ± 13 μmol/L, p=0.021) and at 48 to 72 hours (94 ± 47 versus 39 ± 8 μmol/L, p=0.016).

The brain MRI injury scores are shown in Table 5. The total brain injury score was similar across the groups (8.6 ± 9.7 versus 10.6 ± 10.0, p=0.427). There was no significant difference in the total injury score or the subscores for basal ganglia/thalami, basal ganglia/thalami and proton magnetic resonance spectroscopy (H-MRS), white matter, cerebellar, and additional (intraventricular, subdural hemorrhage, cerebral venous sinus thrombosis) injury between the control and RIC groups.

## DISCUSSION

To our knowledge, this is the first clinical trial evaluating the safety and feasibility of RIC in neonates with HIE undergoing TH. We found that the administration of escalating duration and frequency of RIC during TH was both safe and feasible. Importantly, it was possible to consent caregivers and administer RIC within 24 hours of TH initiation. There were no significant adverse events associated with RIC, including increased pain, vascular, cutaneous, muscular or neural injury, nor were there any differences in seizure burden or mortality. This study was not powered or designed to measure efficacy, but our results provide a critical foundation for the design and implementation of future trials of RIC concomitantly with TH.

RIC has been extensively studied in adults in the context of acute ischemic stroke, where clinical trials have evaluated both pre-hospital and hospital-based RIC interventions (6,10). Evidence for efficacy of RIC in improving neurologic outcome after stroke has been mixed, but in the largest trial of RIC for ischemic stroke, RIC was associated with a greater likelihood of excellent functional outcome at 90 days after symptom onset (6). RIC duration and frequency, as well as patient selection, are key determinants of variability observed in neuroprotection.

RIC likely exerts neuroprotection through several overlapping mechanisms involving neuronal, humoral, and systemic effects (11,12). In the neuronal pathway, the activation of afferent nerves and the release of neuropeptides at the site of RIC act on the efferent nerves at the larger site of ischemia to offer protection (11). The protective effects appear to diminish with the administration of ganglion blockades and nerve resections (13–15). The humoral pathway involves the release of hydrophobic factors that travel from the site of RIC to the site of ischemia for protection (11,12). While the exact constitution of factors remains largely unknown, adenosine, bradykinin-2, and opioids are potential mediators of RIC neuroprotection (11,16–18). Recently, preclinical evidence suggests that two key humoral factors – glucagon-like peptides (GLP) and stromal cell derived factor-1a (SDF-1a) – may be important effectors of RIC organ protection. Both GLP and SDF-1a may increase neurogenesis, reduce cell death, and dampen inflammation (19–22). Importantly, the systemic pathway of RIC benefit likely also involves the modification of the immune system in response to RIC (11), as induction of a homeostatic immune state may promote brain repair. Questions to be addressed include the optimal timing of RIC administration related to the hypoxic ischemic injury, optimal duration, and frequency (i.e., the total RIC dose) for infants in the first days of their HIE.

The main limitation of our study is the small sample size due to its design as a safety and feasibility trial. As such, the current analyses of seizure burden, neuroimaging, and mortality are unable to discern potential immediate benefits of RIC. Future studies with larger cohorts will be needed to determine the potential efficacy of RIC to reduce neurodevelopmental impairment. To enhance recruitment into such future studies, efforts for provider education are required to improve the generalizability of RIC for HIE, as are efforts to improve evidence dissemination and family engagement through patient advocacy groups.

## Conclusion

In this pilot randomized control trial, we found that RIC is safe and feasible in neonates with HIE undergoing TH. Future studies are necessary to investigate the efficacy of RIC in reducing brain injury and improving neurodevelopmental outcomes after HIE.

## Data Availability

All data produced in the present study are available upon reasonable request to the authors.

## Acknowledgements

We would like to thank members of the Data Safety and Monitoring Board (Michael Seed, Mjaye Mazwi, and Andrew Helmers) for their contribution.

## References

1. Kurinczuk JJ, White-Koning M, Badawi N. Epidemiology of neonatal encephalopathy and hypoxic–ischaemic encephalopathy. Early Human Development [Internet]. 2010 Jun 1 [cited 2022 Aug 17];86(6):329–38. Available from: https://www.sciencedirect.com/science/article/pii/S0378378210001088

2. Wu YW, Comstock BA, Gonzalez FF, Mayock DE, Goodman AM, Maitre NL, et al. Trial of Erythropoietin for Hypoxic-Ischemic Encephalopathy in Newborns. N Engl J Med. 2022 Jul 14;387(2):148–59.

3. Azzopardi DV, Strohm B, Edwards AD, Dyet L, Halliday HL, Juszczak E, et al. Moderate Hypothermia to Treat Perinatal Asphyxial Encephalopathy. N Engl J Med [Internet]. 2009 Oct [cited 2022 Aug 17];361(14):1349–58. Available from: https://www-nejm-org.myaccess.library.utoronto.ca/doi/10.1056/NEJMoa0900854

4. Zhao W, Zhang J, Sadowsky MG, Meng R, Ding Y, Ji X. Remote ischaemic conditioning for preventing and treating ischaemic stroke. Cochrane Stroke Group, editor. Cochrane Database of Systematic Reviews [Internet]. 2018 Jul 5 [cited 2022 Sep 28];2019(9). Available from: http://doi.wiley.com/10.1002/14651858.CD012503.pub2

5. England TJ, Hedstrom A, O’Sullivan S, Donnelly R, Barrett DA, Sarmad S, et al. RECAST (Remote Ischemic Conditioning After Stroke Trial). Stroke [Internet]. 2017 May [cited 2022 Sep 28];48(5):1412–5. Available from: https://www.ahajournals.org/doi/10.1161/strokeaha.116.016429

6. Chen HS, Cui Y, Li XQ, Wang XH, Ma YT, Zhao Y, et al. Effect of Remote Ischemic Conditioning vs Usual Care on Neurologic Function in Patients With Acute Moderate Ischemic Stroke: The RICAMIS Randomized Clinical Trial. JAMA [Internet]. 2022 Aug 16 [cited 2022 Sep 28];328(7):627–36. Available from: 10.1001/jama.2022.13123

7. Weir P, Maguire R, O’Sullivan SE, England TJ. A meta-analysis of remote ischaemic conditioning in experimental stroke. J Cereb Blood Flow Metab. 2021 Jan;41(1):3–13.

8. Zozaya C, Ganji N, Li B, Janssen Lok M, Lee C, Koike Y, et al. Remote ischaemic conditioning in necrotising enterocolitis: a phase I feasibility and safety study. Arch Dis Child Fetal Neonatal Ed. 2023 Jan;108(1):69–76.

9. Weeke LC, Groenendaal F, Mudigonda K, Blennow M, Lequin MH, Meiners LC, et al. A Novel Magnetic Resonance Imaging Score Predicts Neurodevelopmental Outcome After Perinatal Asphyxia and Therapeutic Hypothermia. J Pediatr. 2018 Jan;192:33-40.e2.

10. Blauenfeldt RA, Hjort N, Valentin JB, Homburg AM, Modrau B, Sandal BF, et al. Remote Ischemic Conditioning for Acute Stroke: The RESIST Randomized Clinical Trial. JAMA. 2023 Oct 3;330(13):1236–46.

11. Lim SY, Hausenloy DJ. Remote Ischemic Conditioning: From Bench to Bedside. Front Physiol [Internet]. 2012 Feb 20 [cited 2022 Aug 18];3:27. Available from: https://www.ncbi.nlm.nih.gov/pmc/articles/PMC3282534/

12. Hess DC, Khan MB, Hoda N, Morgan JC. Remote ischemic conditioning: a treatment for vascular cognitive impairment. Brain Circ. 2015;1(2):133–9.

13. Schoemaker RG, van Heijningen CL. Bradykinin mediates cardiac preconditioning at a distance. Am J Physiol Heart Circ Physiol. 2000 May;278(5):H1571–1576.

14. Liem DA, Verdouw PD, Ploeg H, Kazim S, Duncker DJ. Sites of action of adenosine in interorgan preconditioning of the heart. Am J Physiol Heart Circ Physiol. 2002 Jul;283(1):H29–37.

15. Dong JH, Liu YX, Ji ES, He RR. [Limb ischemic preconditioning reduces infarct size following myocardial ischemia-reperfusion in rats]. Sheng Li Xue Bao. 2004 Feb 25;56(1):41–6.

16. Tsubota H, Marui A, Esaki J, Bir SC, Ikeda T, Sakata R. Remote postconditioning may attenuate ischaemia-reperfusion injury in the murine hindlimb through adenosine receptor activation. Eur J Vasc Endovasc Surg. 2010 Dec;40(6):804–9.

17. Wolfrum S, Schneider K, Heidbreder M, Nienstedt J, Dominiak P, Dendorfer A. Remote preconditioning protects the heart by activating myocardial PKCepsilon-isoform. Cardiovasc Res. 2002 Aug 15;55(3):583–9.

18. Zhou Y, Fathali N, Lekic T, Ostrowski RP, Chen C, Martin RD, et al. Remote limb ischemic postconditioning protects against neonatal hypoxic-ischemic brain injury in rat pups by the opioid receptor/Akt pathway. Stroke. 2011 Feb;42(2):439–44.

19. Erbil D, Eren CY, Demirel C, Küçüker MU, Solaroğlu I, Eser HY. GLP-1’s role in neuroprotection: a systematic review. Brain Inj. 2019;33(6):734–819.

20. Shan Y, Tan S, Lin Y, Liao S, Zhang B, Chen X, et al. The glucagon-like peptide-1 receptor agonist reduces inflammation and blood-brain barrier breakdown in an astrocyte-dependent manner in experimental stroke. J Neuroinflammation. 2019 Nov 28;16(1):242.

21. Imitola J, Raddassi K, Park KI, Mueller FJ, Nieto M, Teng YD, et al. Directed migration of neural stem cells to sites of CNS injury by the stromal cell-derived factor 1alpha/CXC chemokine receptor 4 pathway. Proc Natl Acad Sci U S A. 2004 Dec 28;101(52):18117–22.

22. Itoh T, Satou T, Ishida H, Nishida S, Tsubaki M, Hashimoto S, et al. The relationship between SDF-1alpha/CXCR4 and neural stem cells appearing in damaged area after traumatic brain injury in rats. Neurol Res. 2009 Feb;31(1):90–102.

